# The Choice of Response Alternatives in COVID-19 Social Science Surveys

**DOI:** 10.1101/2022.01.24.22269741

**Authors:** Daniel B. Wright, Sarah M. Wolff, Rusi Jaspal, Julie Barnett, Glynis M. Breakwell

**Author notes:** Correspondence should be sent to. The Imperial Data set is available at https://github.com/YouGov-Data/covid-19-tracker (Jones, 2020). The de-identified data file for the second study are at: https://github.com/dbrookswr/RespAlt/blob/main/WWRespAlt.csv. The code for all the analyses will appear as the final version as a reproducible knitr (Y. Xie, 2013) document on github.

## Abstract

Social science research is key for understanding and for predicting compliance with COVID-19 guidelines, and much of this research relies on survey data. While much focus is on the survey question stems, less is on the response alternatives presented that both constrain responses and convey information about the assumed expectations of the survey designers. The focus here is on the choice of response alternatives for the types of behavioral frequency questions used in many COVID-19 and other health surveys. We examine issues with two types of response alternatives. The first are vague quantifiers, like “rarely” and ‘frequently.” Using data from 30 countries from the Imperial COVID data hub, we show that the interpretation of these vague quantifiers (and their translations) depends on the norms in that country. If the mean amount of hand washing in your country is high, it is likely “frequently” corresponds to a higher numeric value for hand washing than if the mean in your country is low. The second type are precise numeric response alternatives and they can also be problematic. Using a US survey, respondents were randomly allocated to receive either response alternatives where most of the scale corresponds to low frequencies or where most of the scale corresponds to high frequencies. Those given the low frequency set provided lower estimates of the health behaviors. The choice of response alternatives for behavioral frequency questions can affect the estimates of health behaviors. How the response alternatives mold the responses should be taken into account for epidemiological modeling. We conclude with some recommendations for response alternatives for health behavioral frequency questions in surveys.

---

People’s reactions to mask and COVID-19 vaccine regulations lay bare the need for psychological research to complement biological and economic research for effective management of epidemics. The underlying data for much psychological research come from responses to sample surveys. Constructing survey questions that lead to valid, reliable, and fair responses is difficult (Groves et al., 2009). In everyday conversations, people ask questions without constraining others to respond using pre-defined formats with limited options, but in survey conversations this often occurs. This is done to ease the coding of responses and often to get the respondents to translate their complex beliefs into a single value that is more suitable for statistical analyses.

The responses from health surveys are critical for monitoring health related behaviors, evaluating health campaigns, understanding/modeling the spread of disease, and developing public policy (e.g., Gadarian, Goodman, & Pepinsky, 2021). Health behavior data inform resource allocation decisions and provide necessary information to identify target groups for intervention, to track progress, and to evaluate existing strategies (e.g., Lee & Thacker, 2011). The current research was prompted by the COVID-19 epidemic and the realization that one of the main reasons for its level of impact in many countries is people failing to heed guidelines from scientific groups on the efficacy of health related behaviors (e.g., hand washing, mask wearing, vaccines). Epidemiologists use estimates from social surveys to gauge how much people follow these guidelines.

Self-reports are prone to bias and memory errors (Sudman & Bradburn, 1973). In order to understand measurement error within surveys, researchers examine the cognitive processes that occur when respondents answer questions (e.g., Belli, Conrad, & Wright, 2007; Fienberg, Loftus, & Tanur, 1985; Loftus, Fienberg, & Tanur, 1985; Schaeffer & Dykema, 2020; Tourangeau, 2003). The theoretical approach taken here is to assume the survey situation is an artificial conversation, and like other conversations respondents use the information presented to them to interpret the meaning of questions (Schwarz, 1995). For modeling epidemiology, the focus is often on estimates from behavioral frequency questions (Burton & Blair, 1991), like “how often do you wash your hands?” Our focus is on the response alternatives. As these are not part of most everyday conversations, when they are presented in surveys they may stand out. When responding to a survey question, the responses are often ordered, and respondents will see this as a scale composed of words. According to Grice’s maxims (1975, see also Wilson and Sperber, 2012) when people are presented with a question they assume that the information--including that within the prescribed response format--will be accurate and relevant. While the survey is an artificial conversation, respondents may still assume that the scale and words are appropriate for the behavior in the question (e.g., Schwarz, 1995).

> Respondents assume that researchers construct a meaningful scale that reflects appropriate knowledge about the distribution of the behavior. Accordingly, values in the middle range of the scale are assumed to reflect the ‘average’ or ‘typical’ behavior, whereas the extremes of the scale are assumed to correspond to the extremes of the distribution. Schwarz (2010, p. 49)

If respondents believe the information implied by the set of response alternatives is at odds with what they believe, they could assume the response alternatives were not chosen to have the property Schwarz describes. This could occur for many surveys because some online surveys are poor quality and sometimes designers use the same scale for multiple behaviors without concern of whether it is appropriate for each of these behaviors. This lessens the information likely gathered from the survey and may lower respondents’ view of the designers. Alternatively, the scale could affect what respondents think about the behavior and how they respond (see Figure 1). For example, it may change what respondents think about the behavior in the question stem. If a person is unsure of population norms and is presented with a set of response alternatives suggesting the behavior is common, they may come to believe that the behavior is common. Another possibility is that the response alternatives could change what the respondents think the target event is. Wright, Gaskell, and O’Muircheartaigh (1997) describe how this can occur for vague and ambiguous terms. For example, they asked respondents how often their teeth were cleaned either with response alternatives suggesting this meant by a dentist or with response alternatives suggesting this meant by themselves. Which set respondents received affected what respondents thought the target behavior was. This also occurs for vague behaviors like being annoyed or being satisfied (Gaskell, O’Muircheartaigh, & Wright, 1994; Schwarz, Strack, Müller, & Chassein, 1988). For relatively well defined events (e.g., how many cups of coffee you have in a typical day), the definitions should not be greatly affected. If the difference between the population norms implied by the response alternatives and their pre-survey beliefs is great, this might cause them to doubt the applicability of the scale, or to change their normative beliefs.

**Figure 1.**
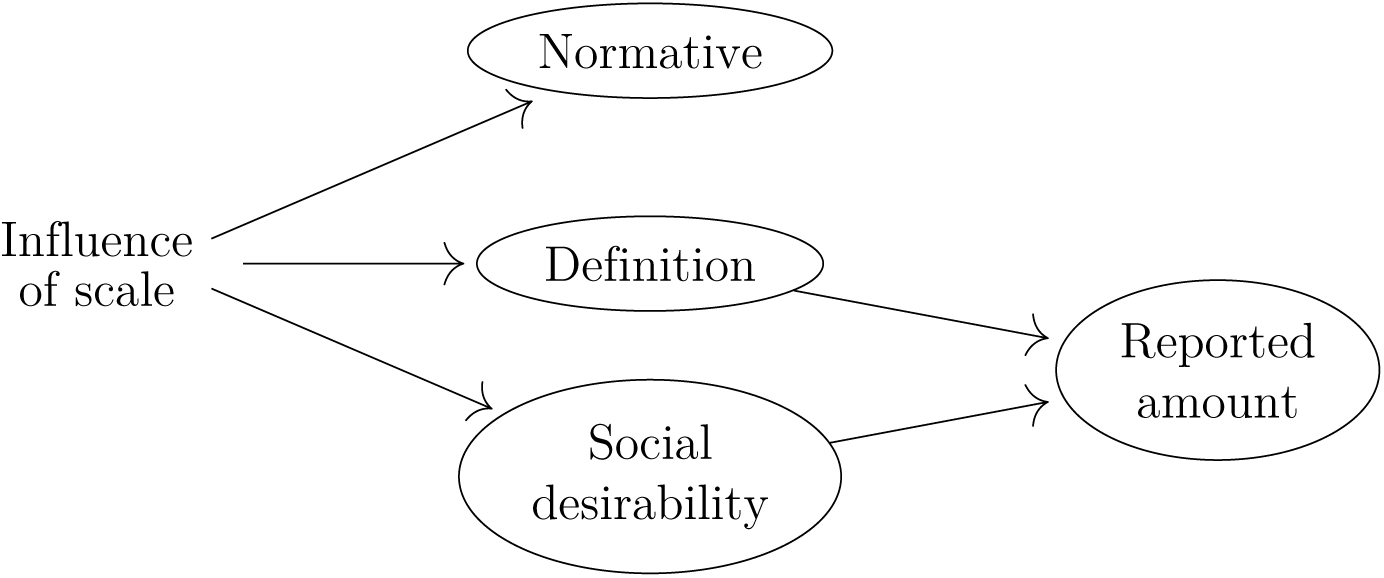
How the response alternatives can affect respondents*’* beliefs about the meaning of the terms, about population norms, and their responses.

Finally, respondents can interpret any set of alternatives as a scale from low to high, ignoring the particular words used to compose the scale. This will be more likely when the response alternatives are vague. In these cases it is unclear what question they answer: how much they engage in the behavior compared with others; with their expectations; with their behavior before the pandemic; etc. This is discussed further at the end of the paper when we make recommendations for the choice of response alternatives. desirability

The response alternatives for behavioral frequency questions generally have one of five formats (more elaborate approaches exist, for example having respondents list the behaviors on a calendar and some of these are discussed in the recommendations section, e.g., Schatz, Knight, Belli, & Mojola, 2020), each with its own concerns:

- <u>Free recall</u>. Respondents provide a numerical estimate, often for a specific duration. This can make difficult memory demands for high frequency behaviors. Respondents often use rough heuristics to *guestimate* the frequency of the behavior. If they try to recall every incident this tends to lead to under-reporting. Another issue is that respondents often round their responses, giving response prototypes (Huttenlocher, Hedges, & Duncan, 1991).
- <u>Last time</u>. Respondents can be asked the last time a relatively infrequent event happened, or if it happened since some memorable event (e.g., Loftus & Marburger, 1983). Time since an event can be used to estimate the event frequency. The difficulty is people have difficulty remembering *when* events occur and tend to forward telescope these dates to be more recent than is accurate (e.g., Neter & Waksberg, 1964; Thompson, Skowronski, & Lee, 1988).
- <u>Comparison</u>. Respondents say how often they experience the behavior compared with others. The difficulty is respondents may not know how often others experience the behavior.
- <u>Vague quantifiers</u>. The respondents use a set of alternatives composed of vague quantifiers to describe how much they experience the behavior. Study 1 examines how different people interpret these differently.
- <u>Numeric response alternatives</u>. Respondents can be provided with a set of exhaustive and mutually exclusive numeric response categories. The concern here, examined in Study 2, is how the choice of these sets can affect how the respondent answers the question and therefore the study’s results.

Before discussing the empirical research, it is important to stress that one of the main limitations of behavioral frequency question is the inaccuracies of human memory. While this is not addressed here, it is important that survey designers take this and other cognitive fallibilities into account when creating surveys.

## Study 1: Vague Quantifiers

Schaeffer (1991) titled her paper “Hardly ever or constantly” as reference to a scene in the film *Annie Hall* where Alvie Singer and Annie Hall are each ask how often they sleep together:

<u>Alvie</u>: Hardly ever, maybe three times a week.

<u>Annie</u>: Constantly, I’d say three times a week.

This highlights that vague quantifiers can be associated with different numeric values for different people.

Wright, Gaskell, and O’Muircheartaigh (1994) showed that part of the differences in how people used vague quantifiers can be attributed to what they think the normative behaviors of that behavior are. They asked UK respondents how much they thought people typical watched televevision, and found large differences in this belief by social class. In a subsequent study they asked the following two questions.

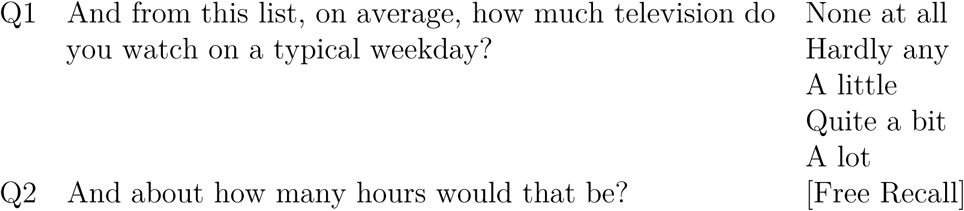

They found that respondents from classes that watched more television and thought the normative behavior was higher believed each vague quantifier corresponded to more hours than did respondents from classes that watched less television. Groups that thought people tended to watch more television interpreted the vague quantifiers as corresponding to higher amounts. Using a large multi-country database, we examine the association between responses using vague quantifiers and a numeric response. Both of these attempt to measure the frequency of the behavior.

Rather than social class, the current study estimates the amount each country does the target behavior and examines if this is a good indicator of how people in the country interpret the vague quantifiers. This study uses data from thirty countries and the survey was delivered in many languages. Linguistic differences make cross-country, and within country where multiple languages are used, difficult. Our prediction is that countries where there is a higher mean for the numeric response, will have higher numeric values corresponding to the different vague quantifiers (and their translations). We split the data into 25% for estimating the means for the behaviors and then used the remaining 75% to test the prediction.

## Methods

Data from https://github.com/YouGov-Data/covid-19-tracker (Jones, 2020) were downloaded on November 8, 2021. The database and discussion of their methods are available at www.coviddatahub.com. In total, at the time of download, there were 734,075 respondents. There are five questions of interest. The first variable is the country. There are thirty countries. Sample sizes of those with complete responses are shown in Table 1.

**Table 1.** Countries in the Imperial database, number of respondents with complete data for the questions analyzed here, and the mean for the natural logarithm of the responses to the free recall question, plus a starting value of +.5, for the 25% of the training sample (given the sample sizes these are very similar to the total means).

| Country | n | mean(ln(x + .5)) | Country | n | mean(ln(x + .5)) |
| --- | --- | --- | --- | --- | --- |
| Australia | 35,932 | 2.03 | Brazil | 10,308 | 2.26 |
| Canada | 31,060 | 2.15 | China | 15,985 | 1.30 |
| Denmark | 32,154 | 2.39 | Emirates | 11,924 | 2.16 |
| Finland | 19,076 | 2.07 | France | 36,360 | 2.07 |
| Germany | 36,271 | 1.93 | Hong Kong | 7,042 | 1.77 |
| India | 16,145 | 2.07 | Indonesia | 12,141 | 1.90 |
| Israel | 6,480 | 1.88 | Italy | 36,106 | 2.19 |
| Japan | 16,931 | 1.49 | Malaysia | 12,133 | 1.91 |
| Mexico | 12,012 | 2.31 | Netherlands | 11,399 | 1.93 |
| Norway | 32,032 | 2.21 | Philippines | 12,002 | 2.07 |
| Saudi Arabia | 11,450 | 1.95 | Singapore | 32,977 | 1.84 |
| South Korea | 15,644 | 1.72 | Spain | 36,198 | 2.06 |
| Sweden | 36,201 | 2.11 | Taiwan | 11,993 | 1.53 |
| Thailand | 12,085 | 1.76 | UK | 46,143 | 2.16 |
| US | 27,919 | 2.02 | Vietnam | 12,074 | 1.75 |

The next three variables of interest are two vague quantifier questions and one free recall question about hand-washing/sanitizing. These were separated by 17 questions. Their wording, from the UK version (i.e., *sanitising*), is shown in Table 2. Only data where there were no missing values for these were used, leaving 646,177 cases. The final question is the Cantril ladder (1965), which asks respondents to imagine a ladder from 0 to 10 steps and asks them to rate their current life satisfaction. About 9% have missing values for this. It is used for exploratory purposes at the end of this section.

**Table 2.** Vague quantifier and free recall hand washing/santizing questions from the Imperial data set.

| Question | Resp. Alts. |
| --- | --- |
| Washed hands with soap and water | Always<br>Frequently<br>Sometimes<br>Rarely<br>Not at all |
| Used hand sanitiser | Always<br>Frequently<br>Sometimes<br>Rarely<br>Not at all |
| Thinking about yesterday ... about how many times, would you say you washed your hands with soap or used hand sanitiser? | [Free recall] |
| Variable names are as found in the Imperial webpage. |  |

## Results and Discussion

The vague quantifier questions are not directly comparable to the free recall question as the latter combines the two behaviors asked about in the vague quantifier questions. We begin by comparing responses from each of vague quantifier questions with the free recall responses question. A small percentage of respondents (0.07%) gave responses of 1,000 or more to the free recall question. Assuming these respondents are awake for 18 hours, this is about once a minute. This is a very skewed variable (skew = 25.06). It was transformed using *ln*(*x* + .5) (the +.5 as some people, 1.22%, said zero) and this lessened the skewness to 0.14.

The associations between the vague quantifier responses and the transformed numeric responses from the free recall question are shown in Figure 2. The relationship between these two variables are fairly weak, even allowing for them asking asking about slightly different behaviors. Twenty-five percent of the data were used to estimate the mean for each country for the transformed (*ln*(*x* + .5)) responses from the free recall question. These are shown in Table 1. The remaining 75% of the data are used to explore the relationship between the transformed numeric responses and the vague quantifiers. Given the large sample size, the 25% and 75% are sufficient for our purposes. Since there are separate vague quantifier questions for hand washing and hand sanitizing, these will be examined separately.

**Figure 2.**
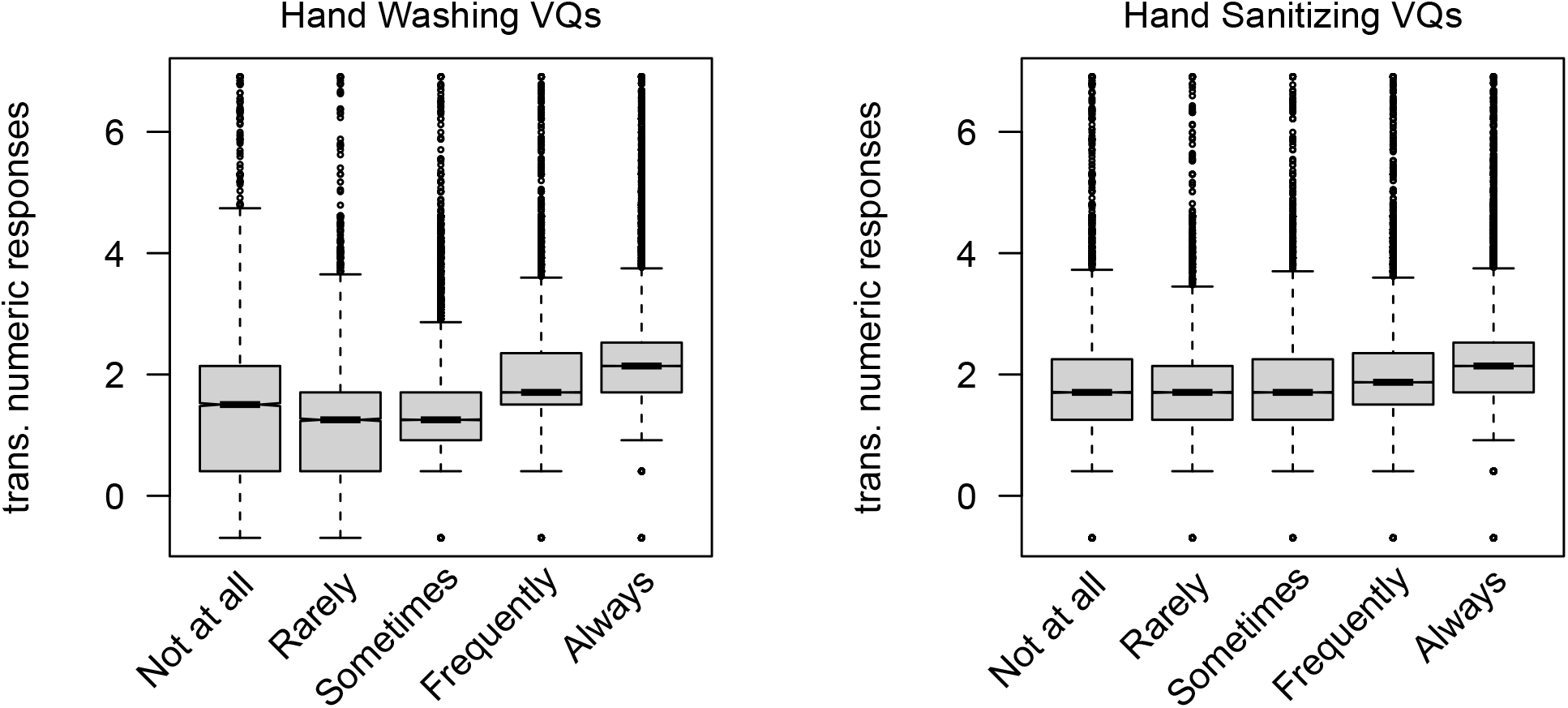
Boxplots for the transformed numeric response from the free recall question for the two vague quantifier questions.

The hand washing vague quantifier variable is treated as categorical, so with *df* = 4 for its five categories. When it is used to predict the transformed variable from the free recall question, the *R*^2^ value was .119. Including the categorical variable of country, with its *df* = 29, increased this to .177. The critical question is how much of this difference, Δ*R*^2^ = .058, can be accounted for by the single variable (df=1) corresponding to the mean of the transformed variable taken from the other 25% of the total sample? If there was 1*/*29th increase the value would be about 3% of this amount, so approximately: .121. It was *R*^2^ = .175, or 96.38% of the possible amount. Figure 3 shows this. The color corresponds to the mean for the transformed free recall responses. The greener the line the lower the country mean for the transformed values (the color is based on a gradient between green and red, so countries with means near the middle appear brown-ish). As is clear, the greener lines are lower than the redder lines.

**Figure 3.**
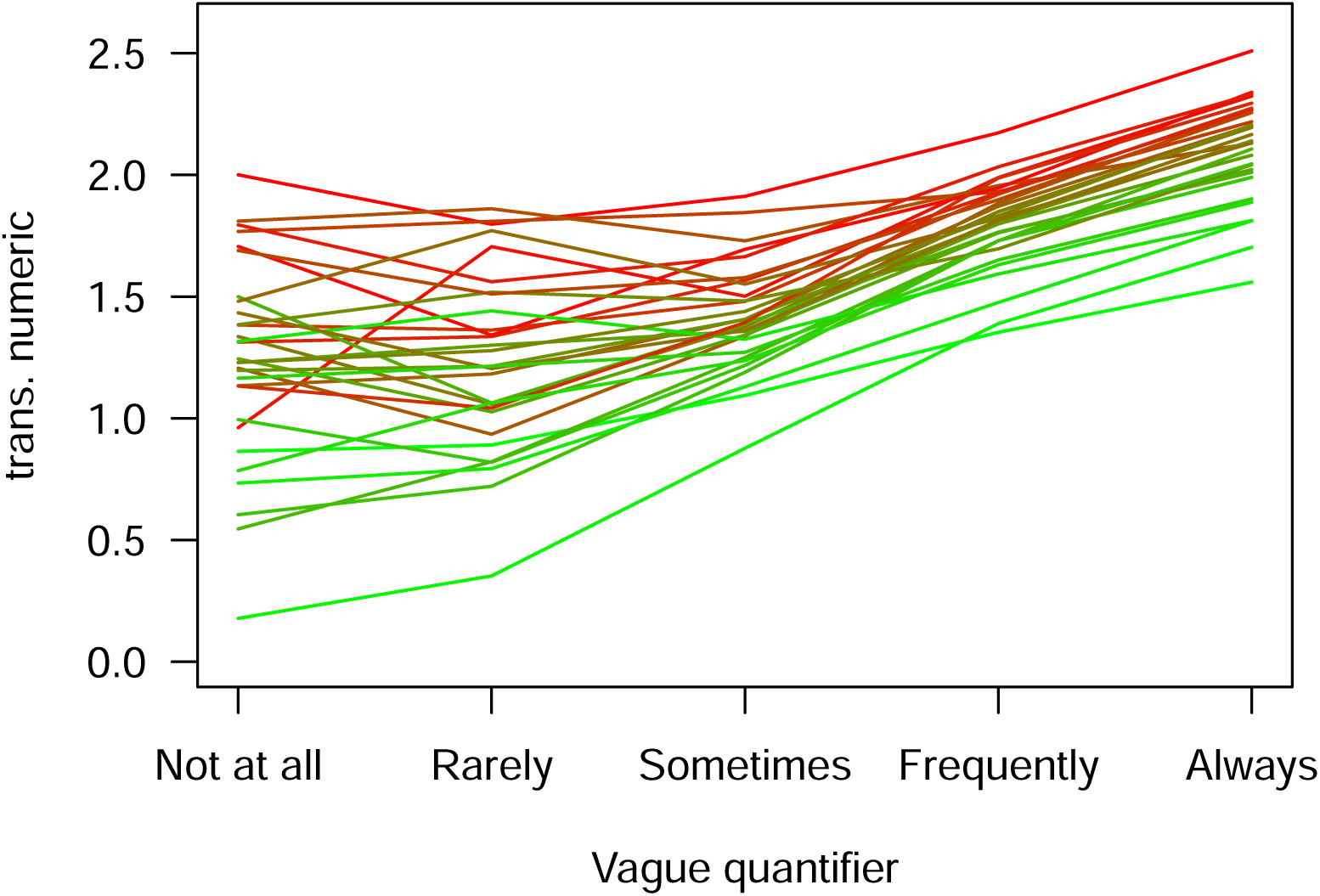
The relationship between the means of the transformed free recall responses for each vague quantifier by country for the vague quantifier washing question. Countries with low free recall responses from the 25% of the sample are shown in **green** and those with high values in **red**, with intermediary countries shown in a mixture of these two colors.

The findings were similar comparing the hand santizing vague quantifier variable with the transformed free recall responses. When just the vague quantifier variable is used to predict the these the *R*^2^ value was .081. Including the categorical variable of country increased this to .143, and difference of Δ*R*^2^ = .058. Using the single country mean variable produced an *R*^2^ = .140, or 93.97% of the possible amount (as opposed to 3%). This is shown in Figure 4 with the greener lines being below the redder lines.

**Figure 4.**
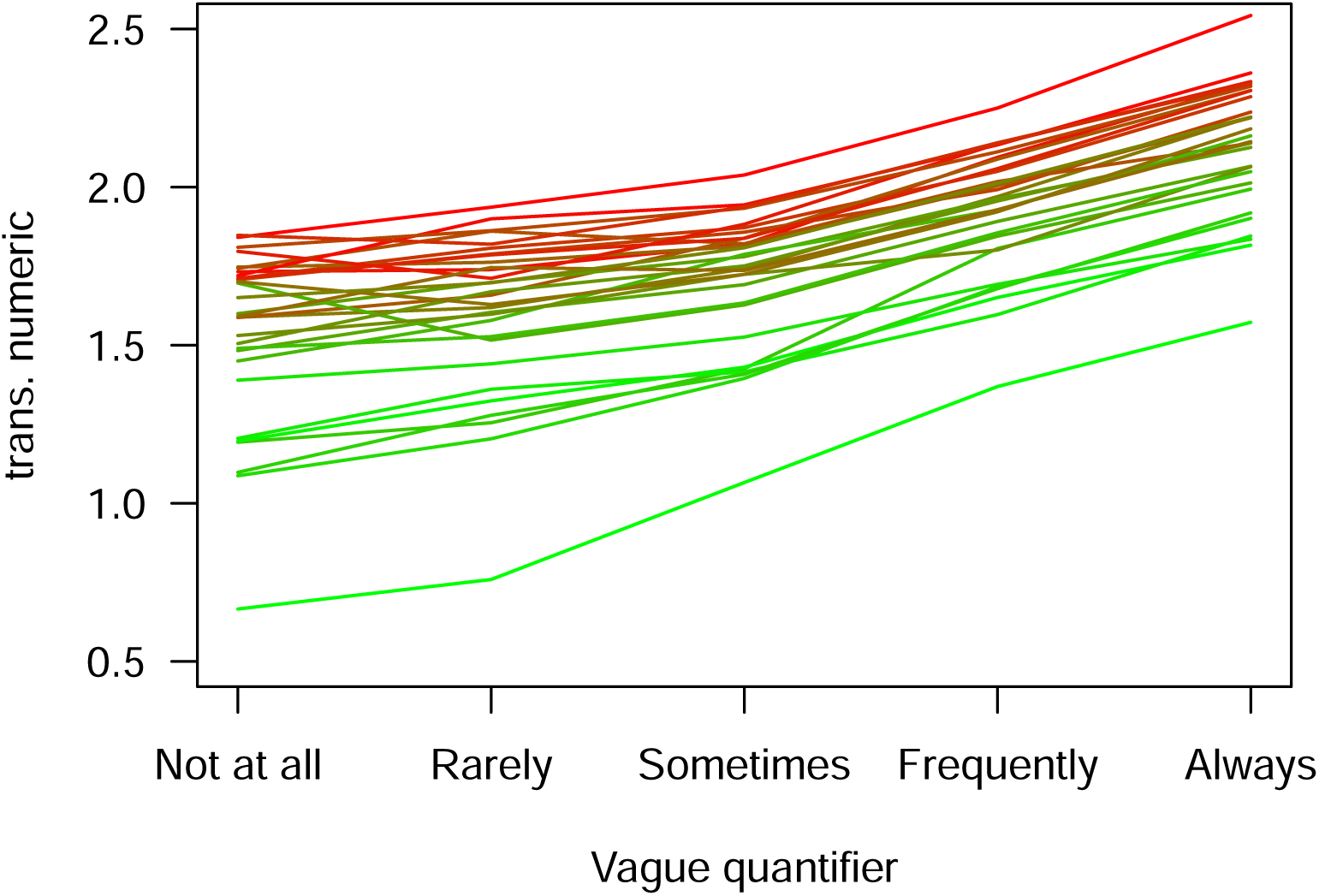
The relationship between the means of the transformed numeric estimates for each vague quantifier by country for the vague quantifier hand sanitizer question. Countries with low free recall responses from the 25% of the sample are shown in **green** and those with high values in **red**, with intermediary countries shown in a mixture of these two colors.

In Study 2 we examine the relationships between response alternatives effects and some attitude questions. Therefore we felt it prudent to examine if an attitude variable, self-expressed life satisfaction (the Cantril Ladder) mediated the relationship between the vague quantifiers and the free recall variable. The Cantril ladder is weakly associated with the transformed free recall responses: Pearson’s *r* = .072. Our interest was whether it has predictive value after accounted for the vague quantifier variables. The *R*^2^ for predicting the transformed free recall variable using the hand washing vague quantifier was *R*^2^ = .123 and using the hand sanitizing vague quantifier was *R*^2^ = .089. Including the interactions between Cantril’s ladder and each vague quantifier question raised these to *R*^2^ = .124 and to *R*^2^ = .090, respectively. These increases are small and will not be considered further.

## Study 2: Numeric Response Alternatives

The purpose of the COVID-19 Household Pulse Survey was to examine the effects of the pandemic on variables ranging from mental health behaviors to vaccine perceptions (US Census Bureau, 2021). The data are meant to help government direct aid, assistance, and support to the people and places that need it most. An example question reads:

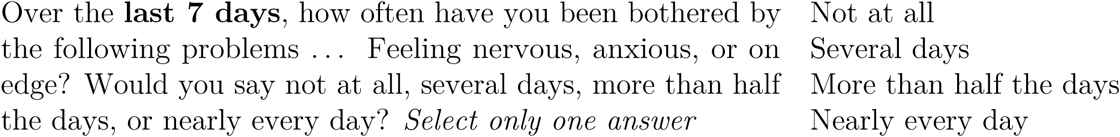

Schwarz and colleagues (Schwarz, 1995; Schwarz & Hippler, 1987; Schwarz, Hippler, Deutsch, & Strack, 1985; Schwarz et al., 1988) conducted a series of studies to show how the choice of which response alternatives to present can affect behavior estimates (see also Gaskell et al., 1994, for use in national surveys). Schwarz uses Grice’s maxims of communication (Grice, 1975) applied to the survey situation explain his findings. Consider the survey question above. The question asks respondents to reflect over the last several days. Three of the four response alternatives involve the event happening multiple days in the previous week: several days; more than half the days; and nearly every day. According to Schwarz this may make respondents feel that being nervous, anxious, or on edge, are likely to occur with greater frequency in the population than if the response alternatives were: more than once a month, once a month, and none.

The behaviors used by Schwarz, Gaskell, and their colleagues were specifically chosen to show the survey methodological effects predicted by Schwarz’s hypotheses. The behaviors used here were chosen because of their relation to disease transmission and that they are part of health guidelines (e.g., those from the Center for Disease Control and Prevention [CDC] in the US, and the National Health Service [NHS] in the UK). Also, Schwarz et al. used in person (face-to-face), pen-and-paper, and telephone administration modes. Nowadays, online surveys are becoming more common so the current study uses the online administration mode. Our primary research question is whether the choice of response alternatives affects the estimates of several health related behaviors. Respondents are randomly allocated into one of two conditions. Those in the first condition are asked behavioral frequency questions with response alternatives that discriminate more finely at low frequencies and those in the second condition are asked these questions with response alternatives that discriminate more finely at high frequencies.

## Methods

The study received IRB approval from the UNLV IRB [1753484-2]. The authors have no conflicts of interest. This study was pre-registered at https://osf.io/b4uef/ and the data are available at https://github.com/dbrookswr/RespAlt/blob/main/WWRespAlt.csv

### Sample

Respondents were recruited via Amazon Mechanical Turk (MTurk). To be an MTurk worker you need to be 18 or over and with US social security number. Additional inclusion restrictions were that respondents had to have 100 previous what are called MTurk human intelligence tasks (HITs) and to have at least a 95% satisfaction rating from those conducting the research. Respondents were compensated $2 for completing the questionnaire.

A *catcha* question was included in the Qualtrics survey. Not correctly completing this meant the survey would not be included, but all passed this (or did not complete the survey). There were two additional exclusion criteria: multiple uses of an IP address and responding too quickly. While each MTurk worker needs a separate US social security number, people could use multiple MTurk accounts. A more likely reason is that people from within the same household are responding using the same IP address. As these people may talk about the study before the second completes it (and would in other ways also be non-independent), the duplicates (i.e., not the first one using the IP address) were excluded. Respondents who on average responded faster than two seconds for the behavioral frequency and attitude questions were also excluded. The number of people excluded for these reasons, in both conditions, is shown in Table 3.

**Table 3.** Respondent flow showing the allocation into conditions and those excluded for a duplicate IP addresses or responding too quickly.

|  | Number of Respondents: Total n = 695 |  |
| --- | --- | --- |
|  | Low Response Alternatives | High Response Alternatives |
| Total Recruited | 345 | 350 |
| Duplicate IP | 16 | 19 |
| Too Quick | 9 | 10 |
| Total excluded | 25 | 29 |
| Percentage excluded | 7% | 8% |
| Final analysed (total n = 641) | 320 | 321 |

### Materials and Experimental Design

Respondents were asked three behavior/health questions related to COVID-19:

- How often did you wash your hands?
- When you washed your hands, typically how long did you spend?
- In a typical day, how often did you apply hand sanitizer?

They were randomly assigned to have either the low or the high response alternatives as listed in Table 4.

**Table 4.**
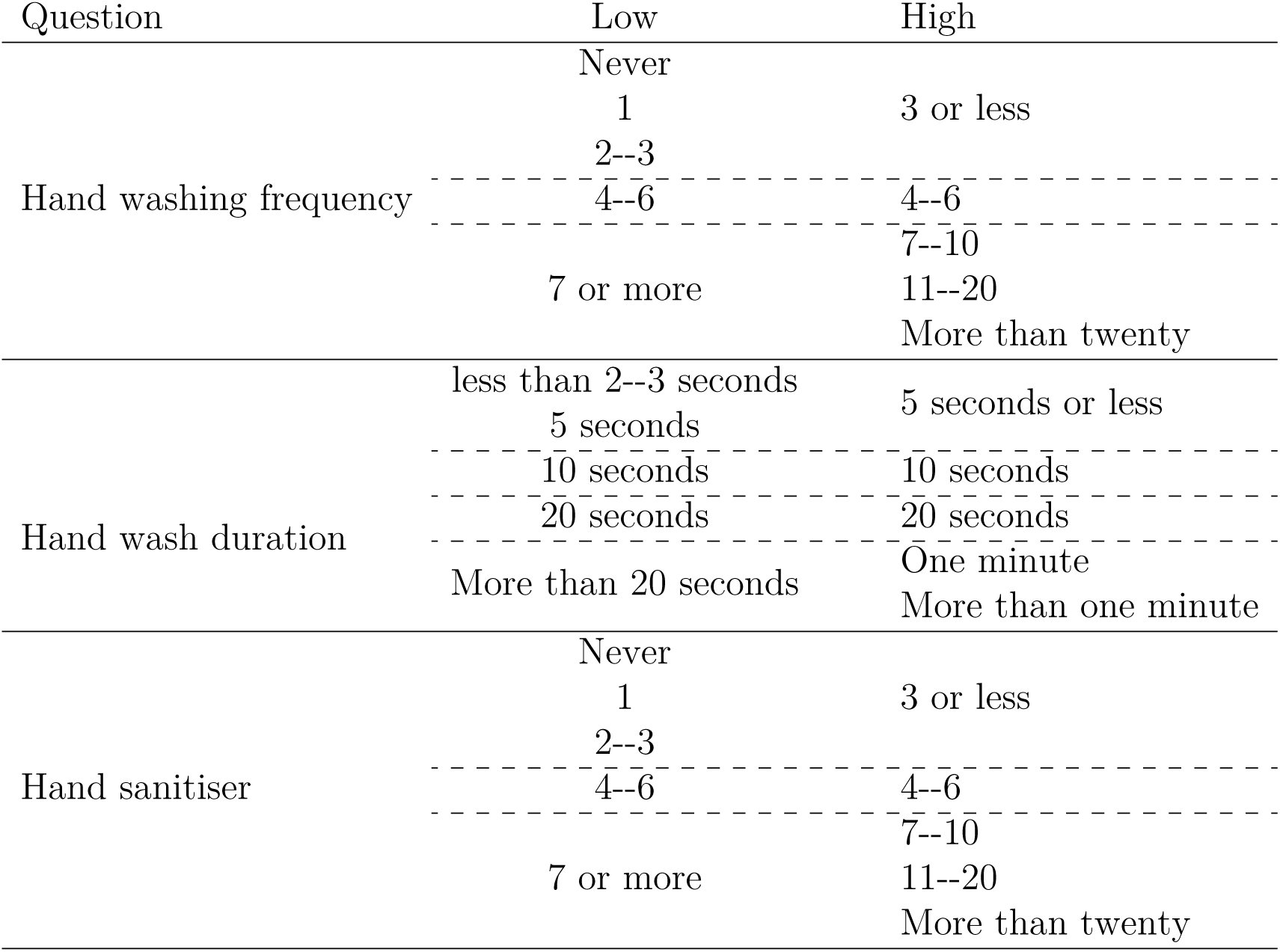
The response alternatives for the low and the high frequency conditions. The dashed lines show how the raw data can be re-coded for comparable frequencies/durations.

Respondents were then asked seven attitude questions are about their health beliefs. Respondents used a 0--100 sliding scale. Responses were measured to the tenth, for example 29.4. Our analyses concerning these variables are exploratory and concern whether the set of response alternatives that were presented affects the responses on these variables.

- Compared with other people, how much were you concerned with the economic impacts of the pandemic?
- Compared with other people, how much were you concerned with the health impacts of the pandemic?
- Thinking back to the previous twelve months, how concerned were you about catching COVID-19 yourself?
- Thinking back to the previous twelve months, how concerned were you about yours friends and family catching COVID-19?
- Suppose that you were supposed to meet a small group of people. If you were not feeling well (slight fever, running nose), how likely would you have stayed at home?
- Suppose that you were not feeling well (slight fever, running nose), how likely is it that you would have consulted a medical professional?
- People vary in how much they trust scientists with respect to many issues. Please rate your view.

In addition, respondents were asked their year of birth, gender, ethnicity, and asked to rate their political beliefs on a 0--100 liberalism/conservativism scale.

The data were collected on a Monday evening, May 17, 2021, and this was a few days after President Biden relayed CDC advice that masks need not be worn by fully vaccinated people indoors in the USA (later, with the increased spread of new variants, guidance changed).

### Statistical Plan

The behavioral frequency questions are treated in two ways. When treating them as 1--5 rating scales, the means of these three are compared between the two groups using Hotelling’s *T* ^2^. When the responses are re-coded into matching semantic categories (within the dashed lines of Table 4), they are treated as categorical variables and compared using *χ*^2^ tests, Cramér’s *V* s, and multinomial logistic regressions. The associations among the attitude questions are examined. The individual item distributions are skewed, so the data are transformed. The associations suggest one underlying dimension. Scores on this dimension are compared for the two groups.

## Results and Discussion

### Behavioral Frequency/Duration Questions

Three related sets of statistical analyses are conducted for examining the behavioral frequency estimates in this study. The first examines if there are differences in responses for the two groups if the response labels are not considered. As such, the questions are all treated as 1--5 rating scales for this set of analyses. The null hypothesis of equal means for the two groups would be true if respondents did not use the response labels. The second set of analyses involves re-coding responses into matching categories, as shown in Table 4. Here the null hypothesis corresponds to respondents not being influenced by whether the response alternatives differentiate more at low or at high frequencies. This is important for epidemiology because if these values differ it would produce different estimates for the frequencies of these behaviors. The third set of analyses is exploratory. It relates to potential moderators of these effects: self-reported political ideology and response time. Whether which set of response alternative is presented affects responses to the attitude questions is examined in the next section.

Figure 5 shows the histograms for the behavior questions when treated as 1--5 rating scales. Table 5 shows the group means, the differences in means, effect sizes for these differences (Cohen’s *d*), and the 95% confidence intervals for these effect sizes. The values of Cohen’s *d* are from 0.65 to 1.11. Cohen (1992) describes 0.5 as a *medium* effect and 0.8 as a *large* effect. Generic verbal labels for effect sizes can be problematic because the absolute meaning of any effect size is context dependent (Baguley, 2004; Lipsey et al., 2012). Here they are used to compare the relative size of these effects with those reported below when the response variables are treated as categorical.

**Figure 5.**
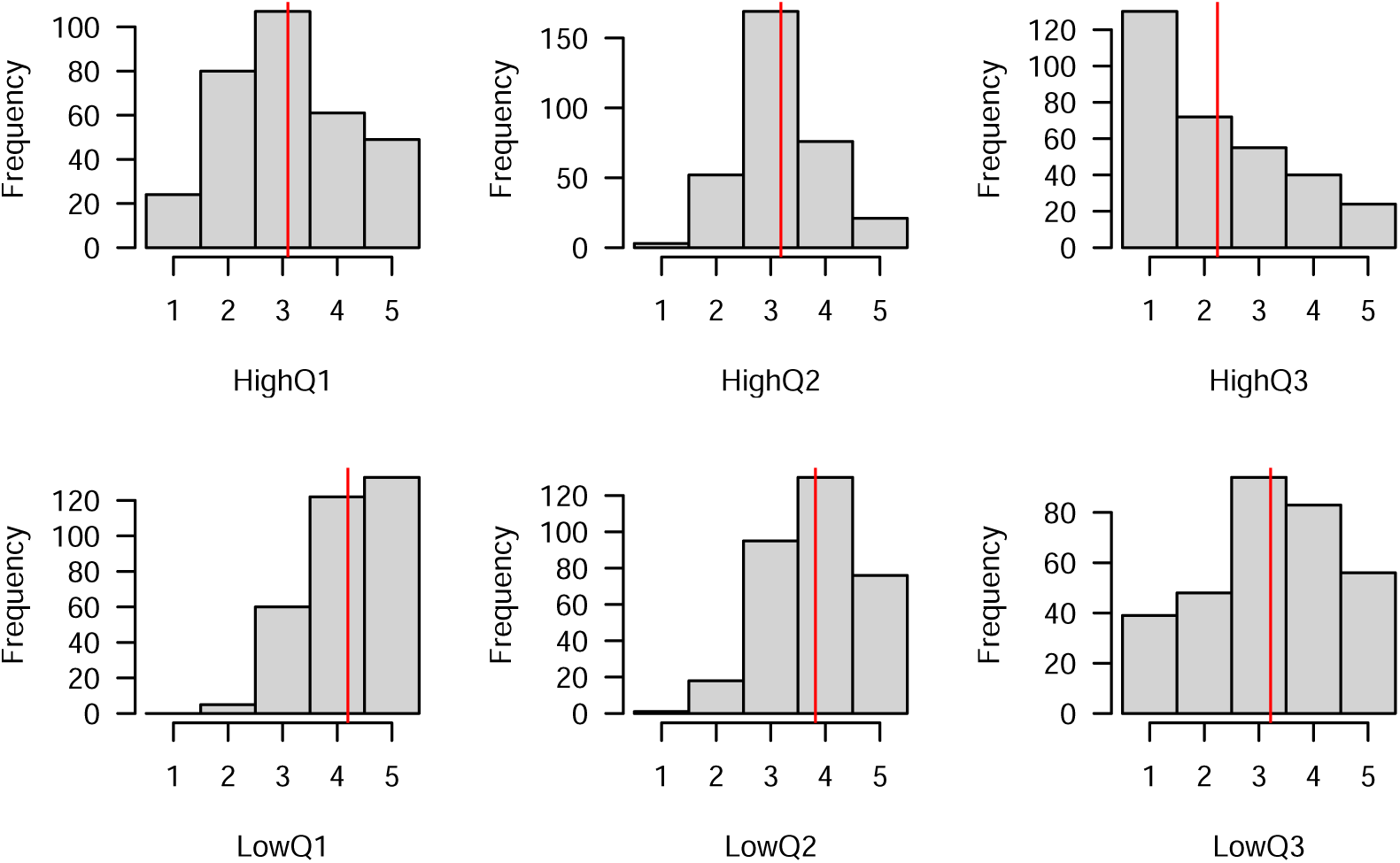
Histograms of the raw behavior responses, by condition. The red shows the condition mean for that variable.

**Table 5.** Descriptive statistics for the raw values, 1--5, for the three behavior answers. This coding ignores the verbal labels, so these differences show some people pay attention to the labels.

| | $\bar{x}_{low}$ | $\bar{x}_{high}$ | diff | LB | UB | Cohen's $d$ | $LB_d$ | $UB_d$ |
| --- | --- | --- | --- | --- | --- | --- | --- | --- |
| Hand wash frequency | 4.20 | 3.10 | -1.10 | 0.95 | 1.25 | 1.11 | 0.94 | 1.27 |
| Hand wash duration | 3.82 | 3.19 | -0.63 | 0.50 | 0.76 | 0.75 | 0.59 | 0.91 |
| Hand sanitizer frequency | 3.22 | 2.24 | -0.98 | 0.78 | 1.17 | 0.75 | 0.59 | 0.91 |

Hotelling’s *T* ^2^ (1931) is used to test if, as a group, the means of these three variables differed by condition. Box’s M (1949), which tests equality of covariances, was statistically significant result, *χ*^2^(6) = 60.38, *p <* .001. Therefore a version of Hotelling’s test that allows for heterogeneity of covariance matrices was used, and (as expected from the effect sizes) it was statistically significant: *F* (3, 619.52) = 236.15, *p <* .001.

The second set of analyses examines the re-coded responses within the dashed lines of Table 4. Table 6 shows the proportions for each of these categories for the two conditions. Cramér’s *V* (with bias correction) is used here as the effect size measure. The BCa bootstrap intervals (2,000 replications) are shown. The *χ*^2^ value is for testing the null hypothesis,

**Table 6.** Statistics comparing the recoded responses for the three behavior questions by condition.

| Question | Cond. | Category | | | | $V$ (95% CI) | $\chi^2(df)$ (all $p < .001$) |
| --- | --- | --- | --- | --- | --- | --- | --- |
|  |  | 1 | 2 | 3 | 4 |  |  |
| Hand wash frequency | Low | .20 | .38 | .42 | | $V = .27$ (.19, .34) | $\chi^2(2) = 47.78$ |
|  | High | .07 | .25 | .68 |  |  |  |
| Hand wash duration | Low | .06 | .30 | .41 | .24 | $V = .22$ (.13, .28) | $\chi^2(3) = 31.85$ |
|  | High | .01 | .16 | .53 | .30 |  |  |
| Hand sanitizer | Low | .57 | .26 | .17 | | $V = .22$ (.14, .29) | $\chi^2(2) = 31.82$ |
|  | High | .40 | .22 | .37 |  |  |  |

*V* = 0. Cohen (1988, p. 222) provides verbal labels for these effect sizes, and they vary by the degrees of freedom. For *df* = 2 (three categories), these are: small = .071, medium = .212, and large = .354. For *df* = 3 (four categories), these are: small = .058, medium = .173, and large = .289. Thus, as a group, using Cohen’s terminology these show medium to large sized effects.

The third set of analyses concerns potential moderators. Two potential moderators of these effects are considered: self-reported political ideology and response time. Two politics variables were used. A political ideology variable is their raw score from 0--100. A political extremeness variable is created, which is the distance from the neutral response of 50, so a 0 to 50 variable. The overall response time variable was skewed, 5.92. The natural logarithm was taken and the skewness was reduced to 0.66. *t*-tests were conducted on all three of these comparing the two conditions. None of these were statistically significant (unadjusted *p*-values shown): for political ideology: *t*(632) = 1.63, *p* = .104; for political extremeness: *t*(632) = 1.81, *p* = .071; and for logged response time: *t*(639) = 0.76, *p* = .450.

Moderation analyses were conducted on the behavior questions when treated as 1--5 rating scales and the re-coded versions. MANOVAs were conducted predicting the three behavior questions as 1--5 rating scales. For each of the three moderator variables, the model with the interaction of the moderator with the condition (high versus low response alternative sets) was compared with the model with just the two main effects. The unadjusted *p*-values were: for political ideology: *p* = .102, for political extremeness: *p* = .809, and for response time: *p* = .889.

The re-coded behavior questions are analyzed individually using multinomial logistic regression (using multinom from the **MASS** package from Venables and Ripley, 2002). The main effect of condition and moderator are included in a model, and this is compared with the model that also includes their interaction. With three behavior questions and three moderators, there are nine *p*-values. The unadjusted values ranged from .026 to .952. Only one of these is lower than the traditional *α* = .05 level when not adjusted for multiple comparisons (those with extreme political views showed a smaller response alternative effect). Applying Holm’s adjustment procedure for multiple comparisons, this becomes *p*_*adjusted*_ = .238. Following Spiegelhalter (2017), the results of exploratory analyses are reported but not elaborated upon.

### Attitude Questions

The attitude questions were included to gauge whether influencing respondents to answer towards one end of the scales would influence how much they reported being worried about COVID-19 compared with others and how extreme their responses on the other attitude questions would be. As discussed in the introduction, the response alternatives may affect respondents about where they believe they place within the population norms. The skewness of the items ranged from -2.45 for seeking medical consultation to -0.54 for staying at home if not feeling well. The variables were transformed by ranking them and normalizing the ranks (see van der Waerden, 1952; Wright, under review). The Pearson correlations of these transformed variables are shown in Table 7.

**Table 7.**
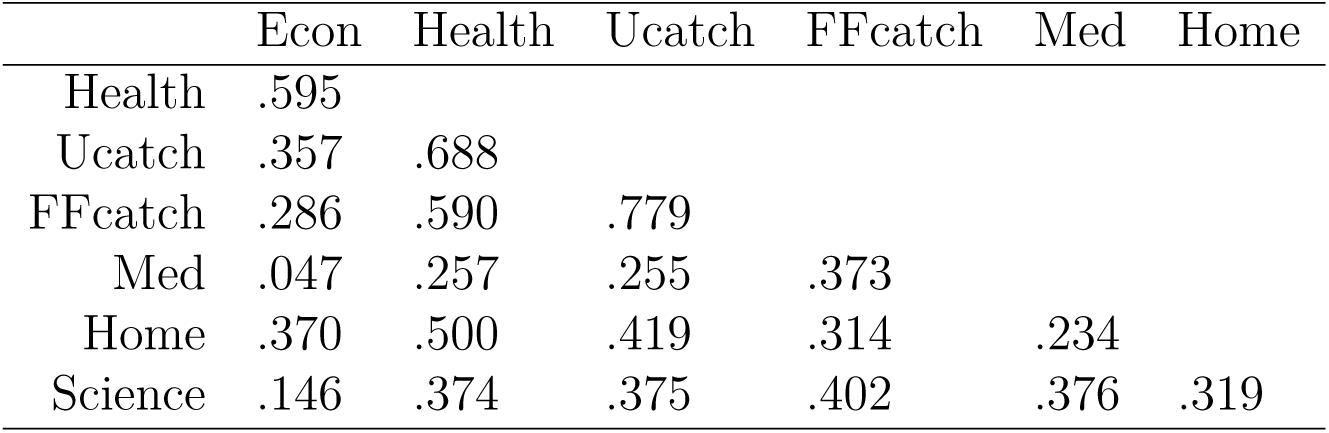
Pearson correlations among the norm-rank transformed attitude questions.

|  | Econ | Health | Ucatch | FFcatch | Med | Home |
| --- | --- | --- | --- | --- | --- | --- |
| Health | .595 |  |  |  |  |  |
| Ucatch | .357 | .688 |  |  |  |  |
| FFcatch | .286 | .590 | .779 |  |  |  |
| Med | .047 | .257 | .255 | .373 |  |  |
| Home | .370 | .500 | .419 | .314 | .234 |  |
| Science | .146 | .374 | .375 | .402 | .376 | .319 |

To increase statistical power, the pre-registered plan was if the items were correlated to combine them into a single dimension. The eigenvalues for the correlation matrix are: 3.41, 1.13, 0.80, 0.62, 0.58, 0.28, and 0.19. Velicer, Eaton, and Fava (2000, see also Auerswald and Moshagen, 2019) discuss how to determine the number of dimensions. A common approach is the empirical Kaiser criterion, which compares the observed eigenvalues with those expected from random data (Braeken & van Assen, 2017). This suggests a single dimension. The first principal component was created and used for comparing groups. The *t*-test comparing means was non-significant: *t*(639) = 0.87, *p* = .382, Cohen’s *d* = 0.07, 95% *CI*_*d*_ = (*-*0.09, 0.22).

## Discussion and Recommendations

Evaluating public health campaigns and modeling disease spread requires estimating people’s behaviors. This is usually done using surveys. Traditionally much effort has focused on the wording of question stems, and less on the response alternatives. Two studies focus on the response alternatives. The first shows that comparing responses across countries using vague quantifiers is difficult, but differences among the countries can almost completely be accounted for by the norms in that country. This provides strong support for the hypothesis put forth in Wright et al. (1994). While this stresses a difficulty making these comparisons, because the country-differences can be accounted for by another variable, a simple theory can be posited:

The meanings of the vague quantifiers are partially based on what respondents believe the survey designers believe about the population norms. Much of the respondents’ beliefs will be based on the norms of their social and national groups.

There will be specific linguistic nuances of particular words and geographic variation in their usage. Further research would be necessary to construct improved ways, based on these differences, to estimate numeric values. However, as noted in the recommendation section, vague quantifiers should not be used if want to estimate or to compare groups with respect to numeric values.

The second study was a randomized experiment to measure the effect of using different response alternatives to estimate the frequency of health related behaviors. The results showed that when several of the response alternatives were for high frequencies, respondents gave answers corresponding to higher estimates than when they were for low frequencies. The choice of response alternatives should be carefully considered when estimating health related behaviors. Further, comparing estimates from surveys that use different sets of response alternatives should be done with great caution, if at all. Which set of response alternatives was presented did not have a statistically significant effect on the attitudinal questions. This suggests that both sets were viewed as reasonably enough by respondents not to create dissonance among them. This is taken into account in our recommendations.

## Recommendations for Response Alternatives

Behavioral frequency questions require respondents to answer questions about past events. We make recommendations for two types of behaviors: rare and frequent. Whether an event type is rare or frequent will depend on the sample and there will be overlap between them, so survey designers should consider all the suggestions below where appropriate.

### Rare events

Consider what is hopefully a rare event for the respondent, like a hospitalization, catching COVID-19, or being laid off. It is likely for most respondents these are rare and it is also likely that answers to these questions will be important both for the survey flow (e.g., on a COVID-19 survey if you answer YES to having COVID-19, you might be asked further questions), and the estimates for these will be important for epidemiological models. Therefore, accurate and precise answers are likely very important. The focus here is on the response alternatives, but it is important to consider the memory limitations, even for rare events (i.e., rare does not imply memorable), and that guiding the respondent using procedures like the cognitive interview, as the term is used in eyewitness research, should be considered (Fisher & Geiselman, 1992). In addition to remembering an event, respondents usually need to provide information about *when* the event occurred. People have difficulty saying when an event occurred (e.g., Friedman, 1993), and this is where issues about the response format are most relevant.

Assuming that the respondents have time to respond to the survey and are near their cell phone, respondents should be encouraged to consult resources (e.g., emails, texts, vaccination card) to provide exact dates. If this information is not available, then respondents could be presented with an *event history calendar*, as devised by Belli, Shay, and Stafford (2001), and used in many health surveys (e.g., Schatz et al., 2020). These allow respondents to fill in notable events, like a child’s graduation and holidays, onto the calendars, and then allow the respondent to think where the event in question happened in relation to these. If using the event history calendar is impractical, most online surveys allow a calendar response so the respondent can give a precise date, and this can also be used if there were multiple incidences in the time frame. For long duration events a start and end could be provided, and respondents could also provide a range if they are uncertain.

### Frequent Events

By frequent events we mean those that, for most respondents, are likely to occur multiple times each week (e.g., handwashing, eating a piece of fruit). As with rare events it is important to consider the cognitive limitations of the respondent, and in particular whether the respondent is likely to use some estimation heuristic or try to recall and count all episodes. Each of these has potential biases (e.g., people are more likely to not remember an event than to create a false memory for a non-existent event, e.g., Wright, Loftus, and Hall, 2001, so recalling each incident is likely to result in an under-estimate), so if the accuracy is of much importance diary methods and experience-sampling methods (e.g., K. Xie, Heddy, & Vongkulluksn, 2019) can be used, though these require coordination with respondents prior to the survey and in the case of experience-sampling methods more technology. For some frequent events, there might be available resources available retrospectively (e.g., filtering through the trash for some food consumption events), though these would not be available for most event types and would be more effort than is likely appropriate. The following assumes these resources are not available and that this is for a single retrospective survey.

Survey designers can be interested in both well-defined events and those that are not well-defined. While standard practice encourages survey questions to be well-defined, if the goal is to compare groups on, for example, how worried they are about catching COVID-19, or some other psychological state, there is often no way to make these precise. Because of this, it will be difficult to interpret numeric estimates of the frequencies with much confidence unless it can be made clear to respondents what, in this case, an episode of worry would be. In these situations it can be prudent to use vague quantifiers. The vagueness of the response alternative matches with the event. Often non-numeric responses can also be used. For example, comparison questions can also be used when this matches what people’s beliefs about the events are like. This would be a situation where pre-testing using think-aloud protocols would likely be valuable to show how respondents think about these event descriptions (Willis, 2005, especially Chapter 4).

For well-defined events, vague quantifiers should be avoided. The choice should be between free recall procedures and a set of numeric response alternatives. We qualify free recall with *ish* to emphasis that this may not mean just writing something in a box. For online surveys, the software can force respondents to enter text is a specified format (e.g., as a number if that is necessary for subsequent analysis, rather than accepting, for example, a respondent writing “about 4 to 8, maybe”). An issue with this is that people’s memories may not be that precise. Goldsmith, Koriat, and Weinberg-Eliezer (2002) describe how people’s memory granularity varies for different memories. This can be accounted for when you ask for numeric information by allowing people to provide a range of values corresponding to their confidence (e.g., Weber & Brewer, 2008). Alternatively, sets of numeric response alternatives can be used, though this has the limitation that the alternatives will usually be a range of values so may not be as precise of free recall methods. However, study 2 showed the the choice of response alternatives can make a difference. We recommend using a large number of alternatives that account for how different groups within the sample will have different expectations.

## Summary

Survey data are used in social science research for informing economists about consumer behaviors, for health researchers evaluating compliance with different campaigns, for sociologists and psychologists constructing theories of why people behave in they ways that they do, and for many other purposes. Behavioral frequency questions have a special place within survey methods as researchers often translate responses into numeric estimates for the behaviors, and sometimes the precision of these estimates is critical (e.g., to epidemiologists predicting trends for the COVID-19 pandemic). While people often focus on the way the event itself is described in the question stem, there is less focus on the response alternatives. We focus on the response alternatives.

Our first study examined how people, across thirty countries, answered questions about hand washing and hand sanitizing. We found that people in different countries interpreted the vague quantifiers used as response alternatives differently. We were able to account for most of the differences among countries using estimates of the behavior in the countries from a different set of respondents. We do not recommend using vague quantifiers with relatively well defined events like hand washing, but allowing people to provide numerical estimates. Our second study showed that care is still necessary when doing this. Using different sets of response alternatives produced different estimates of the behavior.

One consequence of this is that comparisons between studies that use different sets of response alternatives should be done cautiously, if at all. One conclusion from our studies is that the choice of response alternatives should be carefully considered and the deciding how to construct them may be difficult. It may require careful pilot research and techniques like cognitive interviewing and in particular think-aloud protocols (Willis, 2005). We provide a list of recommendations to allow researchers to start thinking about their choices of response alternatives for behavioral frequency questions.

## Data Availability

The complete data set from Imperial College, London, is available at: https://github.com/YouGov-Data/covid-19-tracker The complete de-identified (IP addresses removed) data from the second study are available at: https://github.com/dbrookswr/RespAlt/blob/main/WWRespAlt.csv

https://github.com/dbrookswr/RespAlt/blob/main/WWRespAlt.csv

